# Predicting antibiotic resistance in hospitalized patients by applying machine learning to electronic medical records

**DOI:** 10.1101/2020.06.03.20120535

**Authors:** Ohad Lewin-Epstein, Shoham Baruch, Lilach Hadany, Gideon Y Stein, Uri Obolski

## Abstract

**Background:** Computerized decision support systems are becoming increasingly prevalent with advances in data collection and machine learning algorithms. However, they are scarcely used for empiric antibiotic therapy. Here we accurately predict the antibiotic resistance profiles of bacterial infections of hospitalized patients using machine learning algorithms applied to patients’ electronic medical records.

**Methods:** The data included antibiotic resistance results of bacterial cultures from hospitalized patients, alongside their electronic medical records. Five antibiotics were examined: Ceftazidime (n=2942), Gentamicin (n=4360), Imipenem (n=2235), Ofloxacin (n=3117) and Sulfamethoxazole-Trimethoprim (n=3544). We applied lasso logistic regression, neural networks, gradient boosted trees, and an ensemble combining all three algorithms, to predict antibiotic resistance. Variable influence was gauged by permutation tests and Shapely Additive Explanations analysis.

**Results:** The ensemble model outperformed the separate models and produced accurate predictions on a test set data. When no knowledge regarding the infecting bacterial species was assumed, the ensemble model yielded area under the receiver-operating-characteristic (auROC) scores of 0.73-0.79, for different antibiotics. Including information regarding the bacterial species improved the auROCs to 0.8-0.88. The effects of different variables on the predictions were assessed and found consistent with previously identified risk factors for antibiotic resistance.

**Conclusions:** Our study demonstrates the potential of machine learning models to accurately predict antibiotic resistance of bacterial infections of hospitalized patients. Moreover, we show that rapid information regarding the infecting bacterial species can improve predictions substantially. The implementation of such systems should be seriously considered by clinicians to aid correct empiric therapy and to potentially reduce antibiotic misuse.

**40-word summary:** Machine learning models were applied to large and diverse datasets of medical records of hospitalized patients, to predict antibiotic resistance profiles of bacterial infections. The models achieved high accuracy predictions and interpretable results regarding the drivers of antibiotic resistance.

## Introduction

Antibiotic resistance is a major threat to public health. Substantial increases in antibiotic resistance rates have sprung concerns and bleak estimates as to the future of effective antibiotic treatment [1]. The emergence of antibiotic resistance is mainly shaped by the evolutionary forces of genetic variation (i.e. mutations and horizontal gene transfer) and selection exerted by antibiotic usage. Correspondingly, antibiotic consumption has been repeatedly correlated with increases in antibiotic resistance rates [2]. However, decreases in antibiotic consumption can revert bacterial populations to antibiotic susceptibility, likely due to the fitness cost that antibiotic resistance incurs [3]. Hence, a straightforward intervention to reduce the burden of antibiotic resistance is to decrease antibiotic consumption, for example by reducing inappropriate antibiotic use during empiric therapy [4].

Empiric antibiotic therapy is the commencement of antibiotic therapy before a patient’s precise etiology, source of infection, or antibiotic resistance profile of the infecting pathogen, are confirmed [5]. It is both crucial, as immediate action might be necessary, and by definition based on educated guesses, as it is mostly derived from partial data available to doctors. Two main types of errors occur during empiric therapy – the prescription of inefficient antibiotics (i.e. the antibiotics prescribed do not clear the bacterial pathogen due to its resistance to them), or prescription of antibiotics with too broad of a coverage (i.e. antibiotics with lower coverage would suffice to treat the infection).

The first type of error has more immediate and obvious consequences: treatment with inefficient antibiotics will allow bacteria to keep infecting the patient, putting them at higher risk [6-8]. Furthermore, patients with incorrectly treated infections may be able to keep spreading antibiotic resistance bacteria, causing even greater harm in the future [9]. The second type of error is perhaps not as immediately pronounced but could be detrimental to public health in the long-run. High frequency usage of broad-spectrum antibiotics is likely to increase the frequency of resistance to such antibiotics in the population, as has been observed repeatedly [10-13], rendering these antibiotics less efficient due to accumulated levels of resistance in the population. In turn, this can increase the rate of incorrect empiric therapy of the first kind [14, 15], and lead to increased broad-spectrum antibiotic usage, forming a positive feedback loop of frequent broad-spectrum antibiotic prescription and increased resistance [16, 17]. Moreover, patients treated with broad-spectrum antibiotics can have a substantial part of their microbiome eliminated, enabling subsequent colonization by dangerous and persistent pathogens such as *Clostridium difficile* [18, 19].

A major possible improvement of empiric therapy can stem from using the large amounts of medical data, which are becoming more accessible, in conjunction with machine learning (ML) algorithms. This approach of integrating ML models based on big medical datasets into medical decision making is gaining traction lately and is recognized likely being a part of future treatment in many medical fields [20]. Various studies have identified risk factors for antibiotic resistant infections based on patient comorbidities, demographics, previous received treatments and other patient characteristics [21]. However, identification of risk factors is not necessarily equivalent to highly accurate prediction. Indeed, substantially fewer works have produced models trying to predict antibiotic resistance of infecting bacteria based on patient data. Despite the high quality of many of these studies, they often lacked large datasets [22-24], were limited to a specific types of infection [24-27], pertained only to few bacterial species [23], or only to outpatients [25].

In this work we used electronic medical records of patients hospitalized in Rabin Medical Center, Israel, to predict the antibiotic resistance of bacterial infections. The dataset contained over 16,000 antibiotic resistance tests of bacterial cultures of hospitalized patients with various types of infections, bacterial species, and examined antibiotics. We applied three ML models, and an ensemble combining their results, to predict antibiotic resistance of five antibiotics commonly tested for resistance – Ceftazidime, Gentamicin, Imipenem, Ofloxacin and Sulfamethoxazole-Trimethoprim. We show that accurate antibiotic resistance prediction is possible by using electronic medical records, and that a substantial increase in prediction accuracy occurs if information regarding the infecting bacterial species is available. Finally, we compare the different variables most influencing antibiotic resistance prediction, and explore their effects on resistance probability using two forms of variable influence analysis of the ML models.

## Results

We retrieved medical records of patients which had positive bacterial culture results, from Rabin Medical Center in Israel, from the period between May 2013 and December 2015. The dataset included the bacterial species isolated from the patients and their resistance profiles to the antibiotics tested, as well as the patients’ demographics, comorbidities, hospitalization records and previous antibiotic usage within the hospital (see Methods). In this study we focused on predicting resistance to the five antibiotics most commonly tested for resistance in our dataset: Ceftazidime, Gentamicin, Imipenem, Ofloxacin and Sulfamethoxazole-Trimethoprim (Sul-Trim). All data in the table are aggregated across unique samples.

We split the data into a training set and a test set. The first 85% of each dataset (according to the sample date) were used for training the models while the rest 15% were used for testing the models. We found varying frequencies of antibiotic-resistance, between antibiotics and different bacterial species. The frequencies of antibiotic-resistance also fluctuated through time, yet average resistance frequencies remained similar in the training and test sets (see Figure 1).

**Figure 1.**
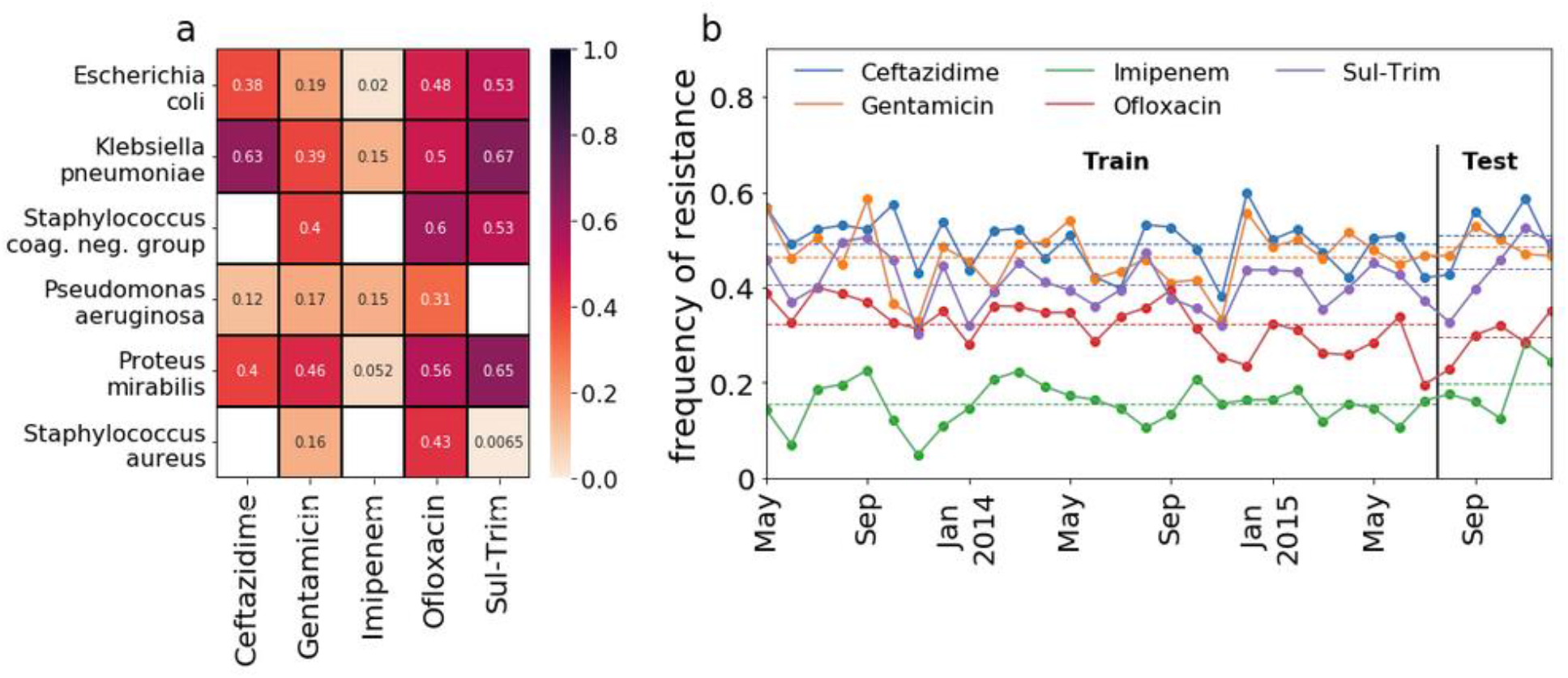
Frequency of antibiotic resistance. **(a)** A heatmap showing the frequencies of antibiotic resistance for each antibiotic and bacterial species combination. Empty cells represent combinations for which there were fewer than 100 data points. **(b)** A time series plot of the frequency of antibiotic resistance observed in each month, for each antibiotic, across all bacterial species. Horizontal dashed lines represent the average resistance frequencies of each antibiotic, separately for the training set and the test set.

We applied ML algorithms to the data in order to generate models for predicting antibiotic resistance of bacterial cultures. We used a supervised ML approach to classify each isolated bacterial culture as either susceptible or resistant to each antibiotic (see Methods and Supplementary Material 2). The final model chosen for predicting antibiotic resistance was an ensemble, composed of three sub-models: L1 regularized logistic regression (LASSO), gradient-boosted decision trees and neural network. Each sub-model was trained separately, and the ensemble provided a prediction based on the average predictions of the sub-models. We examined the success of the ensemble in predicting antibiotic resistance in two data conformations: one where the ensemble was trained and evaluated separately on each antibiotic, and another where the ensemble was trained and evaluated on data containing all five antibiotics, combined. In addition, training and testing of the ensemble was performed once on a dataset that included the identity of the isolated bacterial species, and once on the same data, barring the identity of the isolated bacterial species.

The ensemble achieved high classification success both in terms of area under the receiver-operating-characteristic curve (auROC) and balanced accuracy (i.e., the unweighted average of the sensitivity and specificity rates; see Figures 2 and 3). In addition, the ensemble was found to slightly outperform the sub-models in most scenarios, especially when the identity of the isolated bacterial species was included in the data (see Figure 3).

**Figure 2.**
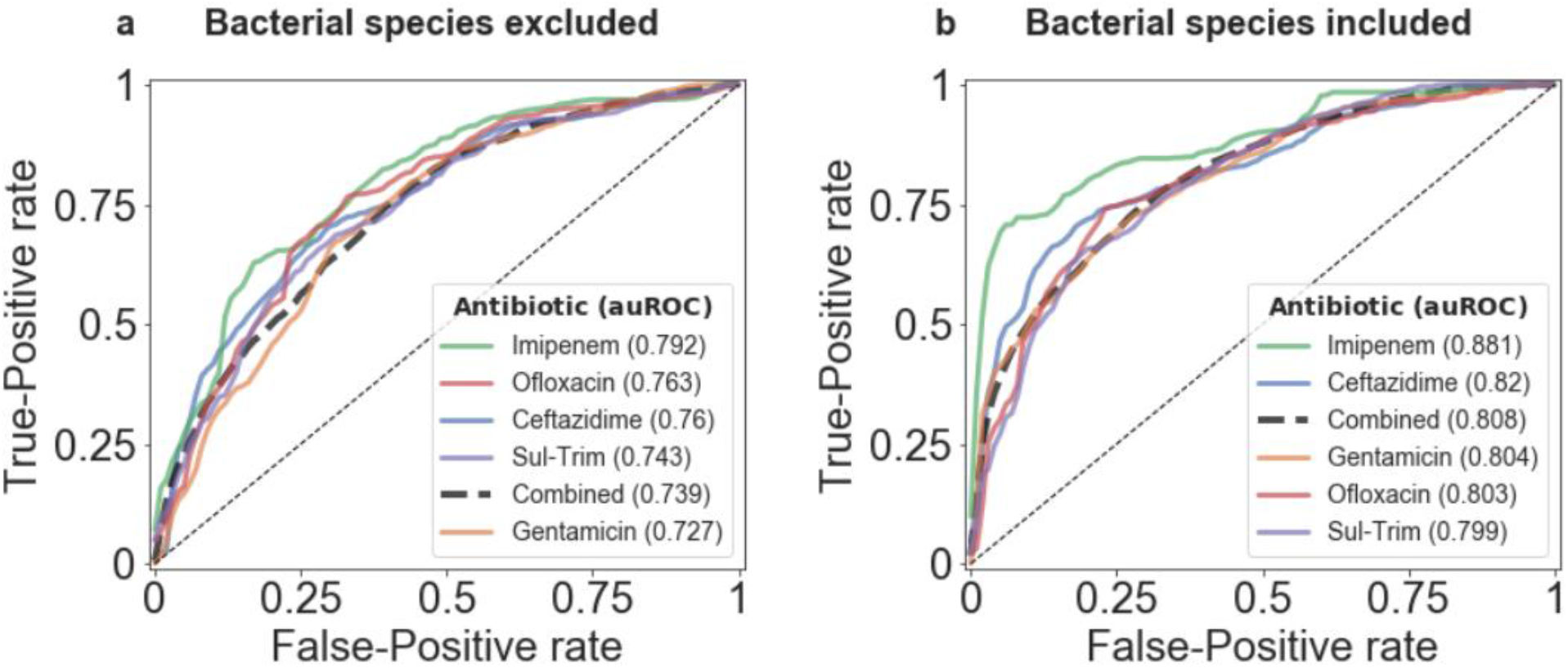
Receiver operating characteristic (ROC) curves of the ensemble. ROC curves of the ensemble are presented seperately for each antibiotic and for all antibiotics combined, for the datasets excluding (a) and including (b) the bacterial species’ identity. The legends show the area under the ROC curve (auROC) for each antibiotic, ordered from highest to lowest.

**Figure 3.**
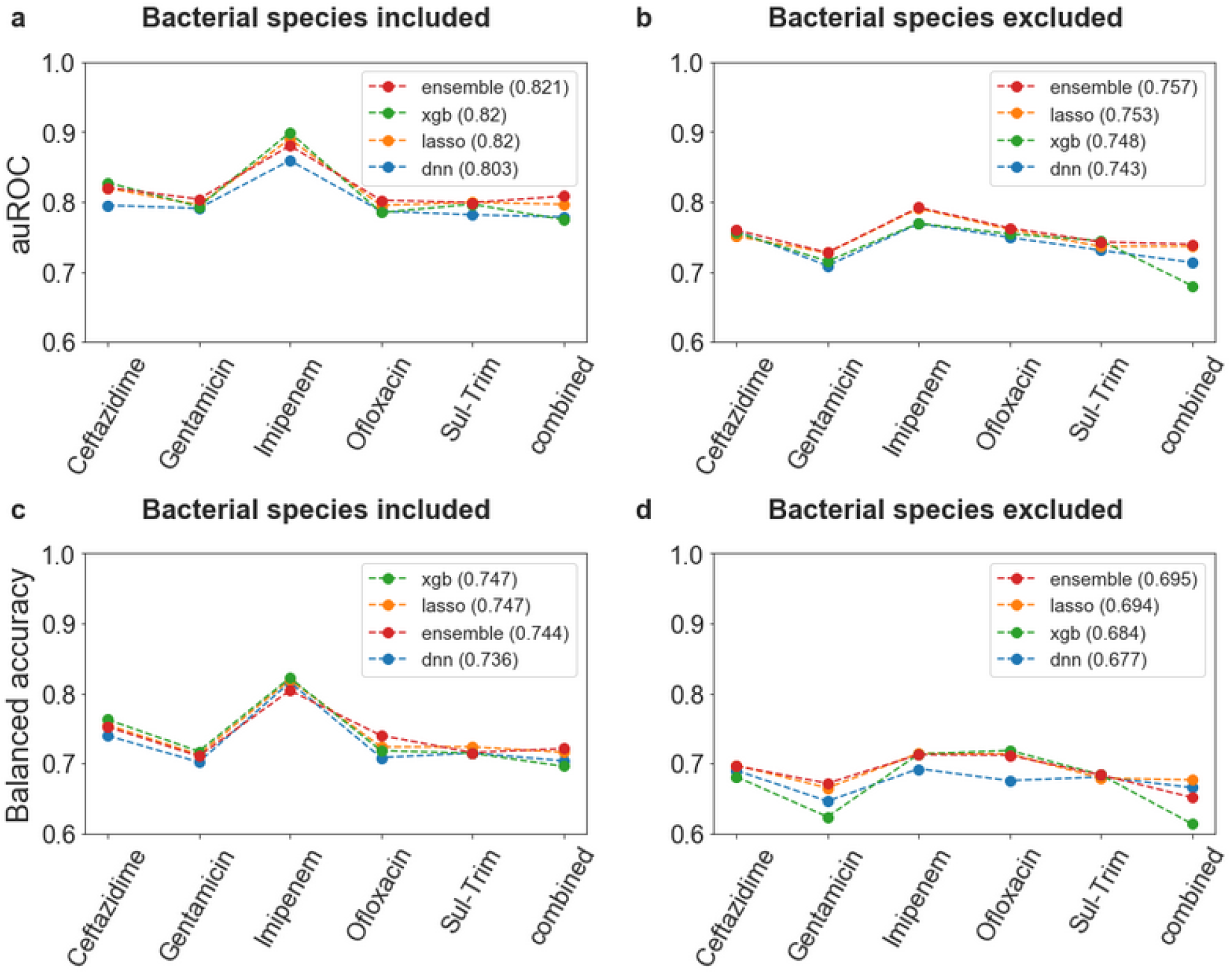
auROC and balanced accuracy scores of the ensemble and its sub-models. The auROC (a, b) and the balanced accuracy (c, d) of the ensemble and its three sub-models, based on data including the identity of the bacterial species (a, c) and on data excluding it (b, d). The legends show the score of each model, averaged over the five antiobiotics, ordered from highest to lowest.

In contrast to classic statistical methods such as regression analysis, the influence of variables on model output is often difficult to gauge in ML models such as boosted trees and neural networks. We thus performed two types of analysis to determine the influence of variables on our ensemble model predictions. First, we performed a permutation-based variable importance analysis (see Methods). Briefly, each variable was randomly permuted to break its association with the outcome. Then, predictions were made using the new dataset with the permuted variable, and the change in the ensemble’s auROC was recorded. Variables for which permutations resulted in substantial decreases in auROC were deemed important. This analysis revealed that the two variables with the highest average effect (across all five antibiotics) were the proportion of past antibiotic resistance infections: previous same-bacterial species resistance to the same antibiotic (previous resistance - specific), and to any antibiotic (previous resistance - general) when including information of the bacterial species; previous any-bacterial species resistance to the same antibiotic (previous any-bacteria resistance - specific), and to any antibiotic (previous any-bacteria resistance - general) when excluding information about the infecting bacterial species (see Supplementary Material 3 and Supplementary tables S1 and S2). Furthermore, we performed a Shapley Additive Explanations (SHAP) analysis [28] (see Methods). The SHAP analysis allowed us to estimate the marginal contribution of each variable to the final prediction of the ensemble. We performed the SHAP analysis separately for each of the five antibiotics tested, both with and without the information regarding the infecting bacterial species. We present the variables having a substantial contribution to prediction of antibiotic resistance (as defined in the Methods) for all five antibiotics in Figure 4. When information regarding the bacterial species was excluded, the two top-contributing variables were consistent with the permutation-based importance analysis - previous any-bacteria resistance - specific and general. These were followed by variables indicating whether the infection was nosocomial or community acquired, and whether the patient was previously treated in the hospital with antibiotics of the same family (antibiotics were categorized into: beta-lactams, fluoroquinolones, aminoglycosides, sulfonamides). Other important variables were the patients’ functioning and independence levels, and previous hospitalization duration. Similarly, when including data regarding the bacterial species, the average previous resistance of the same bacteria to the same/any antibiotic (previous resistance - specific/general, respectively) remained in the top-most affecting variables, alongside indicator-variables of the infecting bacterial species.

**Figure 4.**
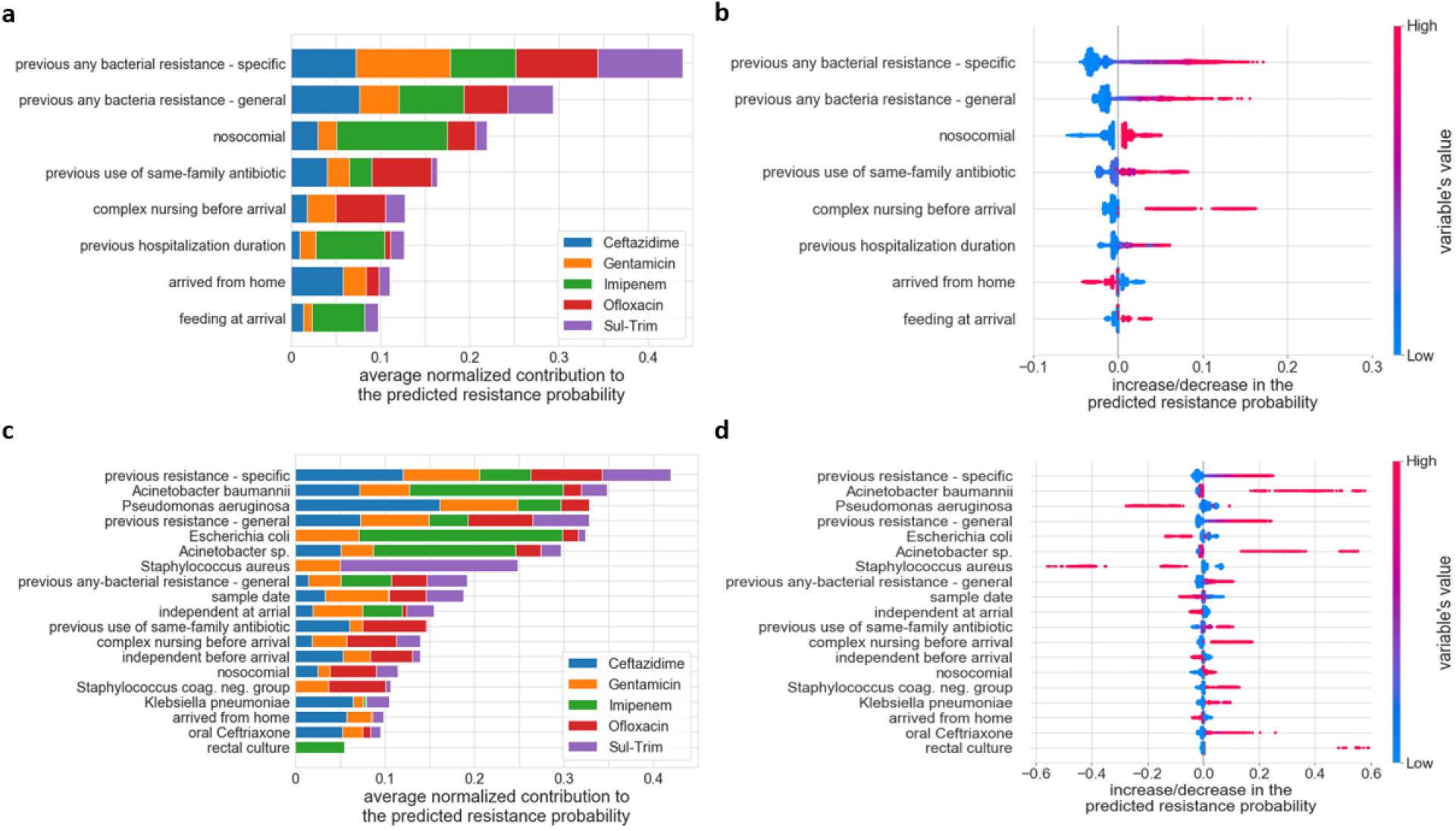
Variable importance analysis using Shapley Additive Explanations (SHAP) **(a, c)** The absolute marginal contribution to predicted probabilities, normalized by the predicted population resistance prevalence (x-axis) is plotted for each antibiotic (color-coded), both for data excluding information on the bacterial species (a) and including it (c). The presented variables are those with an effect of at least 0.05 in any of the five antibiotics. **(b, d)** The marginal changes in predicted resistance probability derived from the same variables shown in (a, c), respectively, are plotted for all antibiotics combined, both for data excluding information on the infected bacterial species (b) and including it (d). Each row in panels (b,d) shows the distribution of the data in two dimensions, and each dot represents one sample: The color represents the value of the variable in a schematic scale from low value to high value (binary variables are represented by the two colors on the edges of the color-bar); the position on the x-axis represents the marginal change in probability of antibiotic resistance due to the variables’ value.

The SHAP analysis also allowed us to investigate whether the different variables in our model act to decrease or increase the probability of antibiotic resistance (Figure 4b,d). Reassuringly, the probability of resistance in our model increased in accordance to known risk factors of antibiotic resistance: previous antibiotic resistant infections, previous hospitalizations, nosocomial infections, previous antibiotic usage, location of sample derivation, and contraindications of patient independence (e.g. nursing home residence and dependence in feeding) [23, 25, 27, 29, 30]. When including information of the infecting bacterial species, additional patterns emerge. For example, while the presence of *Acinetobacter Baumannii* in cultures increases the probability of resistance, *Staphylococcus Aureus* decreases it. The patients’ sex was found to have only a minor effect over the resistance probability, with increased probability of resistance for males. The sample date, which was coded as a numeric variable from the date of the earliest culture in the dataset, was also found to have some effect, probably due to the fluctuating resistance frequencies through time captured by our model.

## Discussion

ML is widely applied in various fields of medicine and is likely to become an invaluable part of medical decision making and treatment [20]. However, ML is scarcely used in aiding the decision of empiric antibiotic therapy. Only a handful of studies have previously utilized the prediction abilities of machine learning models for the rapid detection of antibiotic resistance from patient medical records.

Our work demonstrates the ability to predict antibiotic resistance from patient medical records with high accuracy, and extends previous research in the field in several ways: Rather than relying on a single algorithm, we use an ensemble combining several algorithms substantially differing in their underlying prediction methods (logistic regression, boosted decision trees and neural networks) to produce high-accuracy, robust results. Importantly, we perform a controlled procedure of hyper parameter selection on a training subset of the data and then continue to test our predictions on a disjoint, previously unexplored subset of the data. Furthermore, we predict antibiotic resistance on a large and heterogeneous dataset. It comprises more than 16,000 antibiotic resistance tests of bacterial cultures of hospitalized patients, tested for various antibiotics, and containing multiple bacterial species and infection sites.

Despite the heterogeneity of our data, we were able to train models that achieved highly competitive results: if information regarding the infected bacterial species was excluded we obtained auROC scores in the range of 0.73-0.79, while including the bacterial species yielded an even higher auROC scores in the range of 0.8-0.88. Previous studies which included information regarding the infecting bacterial species obtained auROC scores in the ranges of 0.6-0.83 for antibiotics comparable to those examined in our dataset [27, 31]. Other studies, restricted to one bacterial species or to only one type of infection, had auROC scores in the range of 0.7-0.83 [23-25]. Even when previous auROC results were comparable to those achieved in our study, previous studies did not have such a heterogenic dataset, containing patients with different infections, bacterial species and antibiotics. This added a substantial challenge, which was successfully tackled by our models, and is likely to decrease predictive power of methods used in other studies.

Finally, despite the complex nature of the models used, further complicated by their combination into an ensemble model, we were able to provide interpretation of the influence of different variables on the ensemble’s predictions. Reassuringly, most of the variables found influential in our analysis have been previously identified as increasing risk for antibiotic resistant infections. In addition to further validating our model against prior knowledge, understating which variables are influential can help indicate important drivers of antibiotic resistance. For example, the variables consistently highly ranked as important in our models were those pertaining to previously resistant bacterial cultures. The importance of those variables might imply the persistence of resistant bacterial flora in patients, and may warrant further investigations into treatments restoring the normal bacterial flora after antibiotic treatments, especially in patients at risk for re-hospitalization [32]. However, such conclusions should be further investigated in other settings before concrete conclusions can be reached.

An especially important variable in our models was the identity of the bacterial species causing the infection. It is plausible that inherent biological differences, combined with different exposures to antibiotics, produce the substantial observed differences between resistance frequencies of different bacterial species (Figure 1), and hence rendered this information predictive in our models. Although not routinely performed in most hospitals, very rapid identification of the infecting bacterial species is possible, e.g. through PCR-based methods [33]. If our predictive model is to be implemented in real time in clinical settings, adding such rapid bacterial species identification tools might be cost-beneficial, given the improvement in our prediction results, and the major cost incurred by antibiotic resistant infections [34].

Additional potentially important variables are various community-derived risk factors for antibiotic resistant infections such as antibiotic use outside the hospital [25, 35, 36], patient location of residency [12], and other factors as microbiome composition, diet and exercise [37-40]. Unfortunately, these were not available to us and hence not included in our analysis. Our results can likely be improved by inclusion of such data, and future work should consider their inclusion when available.

To conclude, our results present an ML approach to predict antibiotic resistance of bacterial infections of hospitalized patients, using the patient’s electronic medical record. Our method autonomously identified known risk factors of antibiotic resistance, and provided high-accuracy predictions based on the complex interactions between them and other patient information. Applying our approach, and further developing it by incorporating additional patient data is paramount for achieving highly informed, personalized empiric antibiotic therapy. Such therapy should result in less antibiotic misuse, and hopefully aid the fight against antibiotic resistance.

## Methods

### Data processing and variable engineering

We processed the datasets described in the Results section and in Table 1, and created variables that were fed into the machine learning models. All categorical variables were one-hot encoded as dummy variables. A total of 322-448 variables were included in the processed dataset (depending on the categorical variables in each dataset, and on whether bacterial species information was included). A description of the variables that were concluded most important is available at Supplementary material 1. We considered intermediate resistance results as resistant for both simplicity of classification and under a conservative rationale, as is common practice [25].

**Table 1.**
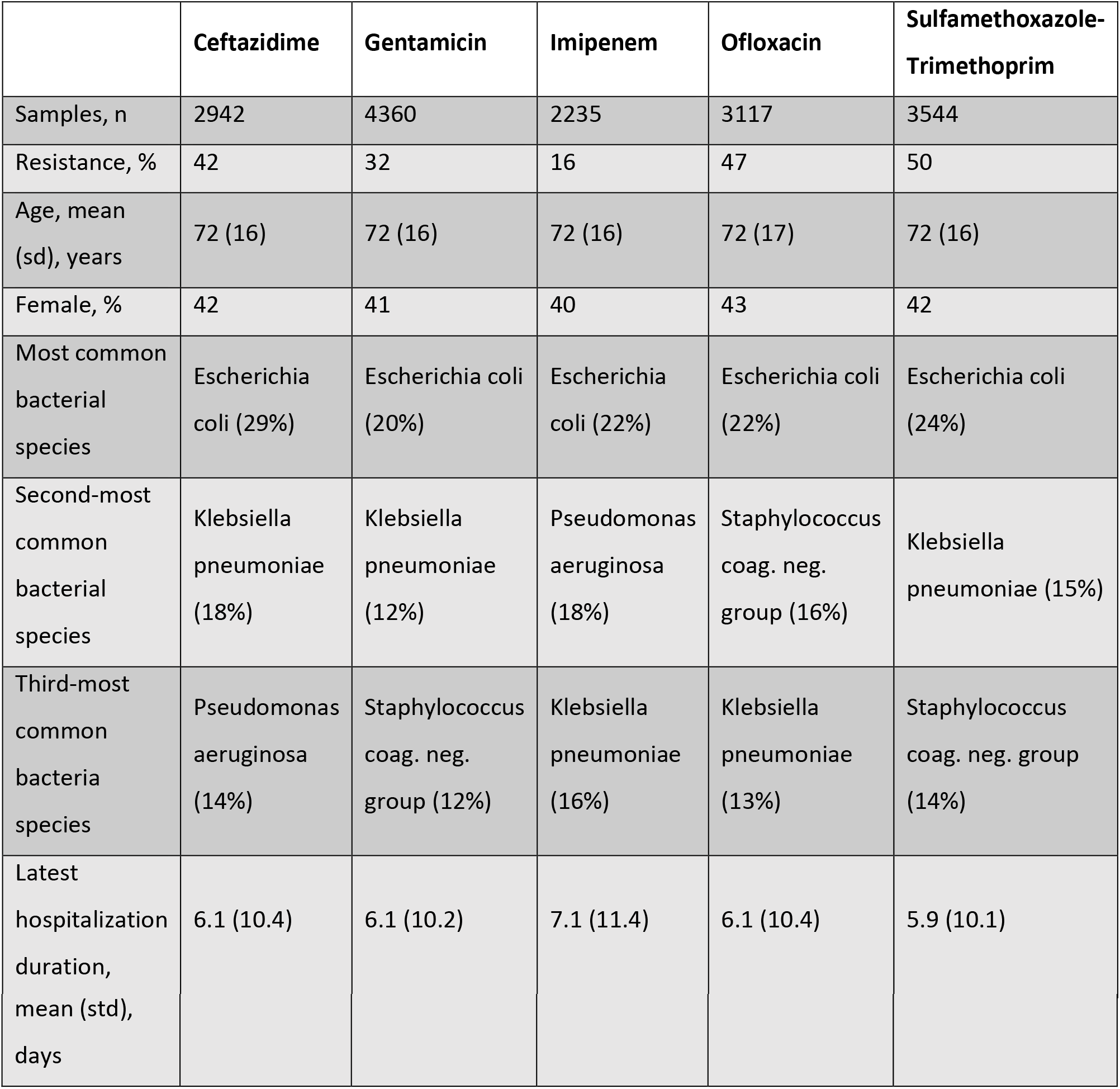
Summary statistics of the dataset.

### Machine learning models

We used an ensemble model, composed of three sub-models: L1 regularized logistic regression (LASSO), gradient-boosted decision trees and neural network. Each sub-model was trained separately on the data and for each data point it provided a prediction: a number in the range 0-1. The ensemble prediction is the average of the predictions of these three models (Supplementary Material 2).

Each data point input into the models contained the abovementioned variables as well as the susceptible or resistant binary label for a single antibiotic as the predicted variable. For each antibiotic, the data were divided to distinct training and test sets based on dates, so that the early 85% of the data were assigned to the training set.

### Model evaluation

After performing the hyperparameter tuning and variable selection on the training set (using cross validation), the chosen models were applied to the test set. The ensemble’s predictions were compared to the actual resistance class of each data point to derive the auROC score.

When computing the balanced accuracy score, we used the first 85% of the training set in order to train the ensemble, and the remaining 15% of the training set (validation set) in order to find the optimized prediction threshold *ρ* ∈ (0,1), under which each prediction was assigned to 1 if above *ρ* and to 0 if below *ρ*. The thresholds were chosen to maximize the balanced accuracy score on the validation set. We then trained the ensemble on the entire training set and used it to predict results on the test set. We then compared the ensemble’s predictions to the actual class of each data point, and derived the balanced accuracy score.

### Variable importance analysis - permutation tests

Each variable’s values were randomly permuted in the test set, while other variables kept as they were. After a permutation was performed, the auROC of the ensemble’s prediction on the test set was re-calculated. The absolute difference of the resulting auROC from that obtained on the original test set was recorded. This was repeated 100 times (for each variable) and the average result was deemed as each variable’s importance score.

### Variable influence analysis - Shapley Additive Explanations (SHAP)

We applied the SHAP analysis to the training sets, performing the analysis separately for each sub-model, and then averaging the results to obtain the ensemble’s scores (supplementary material 3). The SHAP values can be interpreted as the marginal change in the probability of resistance of each observation, derived for each variable.

### Software used

All analyses were performed using Python 3.6.

## Data Availability

Patient data is not made available. Data regarding the analysis is fully available in the supplementary material.

## Acknowledgements

We gratefully acknowledge the support of NVIDIA Corporation with the donation of the Titan Xp GPU used for this research.

## References

1. Ventola CL. The antibiotic resistance crisis: part 1: causes and threats. Pharmacy and therapeutics 2015; 40(4): 277.

2. Bell BG, Schellevis F, Stobberingh E, Goossens H, Pringle M. A systematic review and meta-analysis of the effects of antibiotic consumption on antibiotic resistance. BMC infectious diseases 2014; 14(1): 13.

3. Melnyk AH, Wong A, Kassen R. The fitness costs of antibiotic resistance mutations. Evolutionary applications 2015; 8(3): 273–83.

4. Laxminarayan R, Duse A, Wattal C, et al. Antibiotic resistance—the need for global solutions. The Lancet infectious diseases 2013; 13(12): 1057–98.

5. Mandell G, Dolin R, Bennett J. Mandell, Douglas, and Bennett’s principles and practice of infectious diseases: Elsevier, 2009.

6. Paul M, Shani V, Muchtar E, Kariv G, Robenshtok E, Leibovici L. Systematic review and meta-analysis of the efficacy of appropriate empiric antibiotic therapy for sepsis. Antimicrobial agents and chemotherapy 2010; 54(11): 4851–63.

7. Oshima T, Kodama Y, Takahashi W, et al. Empiric antibiotic therapy for severe sepsis and septic shock. Surgical infections 2016; 17(2): 210–6.

8. Ibrahim EH, Sherman G, Ward S, Fraser VJ, Kollef MH. The influence of inadequate antimicrobial treatment of bloodstream infections on patient outcomes in the ICU setting. Chest 2000; 118(1): 146–55.

9. Obolski U, Stein GY, Hadany L. Antibiotic restriction might facilitate the emergence of multi-drug resistance. PLoS computational biology 2015; 11(6).

10. Paterson DL. “Collateral damage” from cephalosporin or quinolone antibiotic therapy. Clinical Infectious Diseases 2004; 38(Supplement_4): S341-S5.

11. Vernaz N, Huttner B, Muscionico D, et al. Modelling the impact of antibiotic use on antibiotic-resistant Escherichia coli using population-based data from a large hospital and its surrounding community. Journal of Antimicrobial Chemotherapy 2011; 66(4): 928–35.

12. Low M, Neuberger A, Hooton TM, et al. Association between urinary community-acquired fluoroquinolone-resistant Escherichia coli and neighbourhood antibiotic consumption: a population-based case-control study. The Lancet Infectious Diseases 2019; 19(4): 419–28.

13. Pantosti A, Moro ML. Antibiotic use: the crystal ball for predicting antibiotic resistance. The University of Chicago Press, 2005.

14. Merli M, Lucidi C, Di Gregorio V, et al. The spread of multi drug resistant infections is leading to an increase in the empirical antibiotic treatment failure in cirrhosis: a prospective survey. PLoS One 2015; 10(5).

15. Carrara E, Pfeffer I, Zusman O, Leibovici L, Paul M. Determinants of inappropriate empirical antibiotic treatment: systematic review and meta-analysis. International journal of antimicrobial agents 2018; 51(4): 548–53.

16. Kollef MH. Appropriate empirical antibacterial therapy for nosocomial infections. Drugs 2003; 63(20): 2157–68.

17. Murthy R. Implementation of strategies to control antimicrobial resistance. Chest 2001; 119(2): 405S-11S.

18. Crowther GS, Wilcox MH. Antibiotic therapy and Clostridium difficile infection-primum non nocere-first do no harm. Infection and drug resistance 2015; 8: 333.

19. Fridkin S, Baggs J, Fagan R, et al. Vital signs: improving antibiotic use among hospitalized patients. MMWR Morbidity and mortality weekly report 2014; 63(9): 194.

20. Rajkomar A, Dean J, Kohane I. Machine learning in medicine. New England Journal of Medicine 2019; 380(14): 1347–58.

21. Control CfD, Prevention. Antibiotic resistance threats in the United States, 2013: Centres for Disease Control and Prevention, US Department of Health and 2013.

22. Oonsivilai M, Mo Y, Luangasanatip N, et al. Using machine learning to guide targeted and locally-tailored empiric antibiotic prescribing in a children’s hospital in Cambodia. Wellcome open research 2018; 3.

23. Sullivan T, Ichikawa O, Dudley J, Li L, Aberg J. The rapid prediction of carbapenem resistance in patients with Klebsiella pneumoniae bacteremia using electronic medical record data. In: Open forum infectious diseases: Oxford University Press US, 2018:ofy091.

24. Dan S, Shah A, Justo JA, et al. Prediction of fluoroquinolone resistance in Gram-negative bacteria causing bloodstream infections. Antimicrobial agents and chemotherapy 2016; 60(4): 2265–72.

25. Yelin I, Snitser O, Novich G, et al. Personal clinical history predicts antibiotic resistance of urinary tract infections. Nature medicine 2019; 25(7): 1143–52.

26. Dickstein Y, Geffen Y, Andreassen S, Leibovici L, Paul M. Predicting antibiotic resistance in urinary tract infection patients with prior urine cultures. Antimicrobial agents and chemotherapy 2016; 60(8): 4717–21.

27. Vazquez-Guillamet MC, Vazquez R, Micek ST, Kollef MH. Predicting resistance to piperacillin-tazobactam, cefepime and meropenem in septic patients with bloodstream infection due to Gram-negative bacteria. Clinical Infectious Diseases 2017; 65(10): 1607–14.

28. Lundberg SM, Lee S-I. A unified approach to interpreting model predictions. In: Advances in neural information processing systems, 2017:4765-74.

29. Chatterjee A, Modarai M, Naylor NR, et al. Quantifying drivers of antibiotic resistance in humans: a systematic review. The Lancet Infectious Diseases 2018; 18(12): e368-e78.

30. MacFadden D, Coburn B, Shah N, et al. Utility of prior cultures in predicting antibiotic resistance of bloodstream infections due to Gram-negative pathogens: a multicentre observational cohort study. Clinical Microbiology and Infection 2018; 24(5): 493–9.

31. Tandan M, Timilsina M, Cormican M, Vellinga A. Role of patient descriptors in predicting antimicrobial resistance in urinary tract infections using a decision tree approach: A retrospective cohort study. International journal of medical informatics 2019; 127: 127-33.

32. Francino M. Antibiotics and the human gut microbiome: dysbioses and accumulation of resistances. Frontiers in microbiology 2016; 6: 1543.

33. Järvinen A-K, Laakso S, Piiparinen P, et al. Rapid identification of bacterial pathogens using a PCR-and microarray-based assay. BMC microbiology 2009; 9(1): 161.

34. Dadgostar P. Antimicrobial Resistance: Implications and Costs. Infection and Drug Resistance 2019; 12: 3903.

35. Costelloe C, Metcalfe C, Lovering A, Mant D, Hay AD. Effect of antibiotic prescribing in primary care on antimicrobial resistance in individual patients: systematic review and meta-analysis. Bmj 2010; 340: c2096.

36. Pouwels KB, Freeman R, Muller-Pebody B, et al. Association between use of different antibiotics and trimethoprim resistance: going beyond the obvious crude association. Journal of Antimicrobial Chemotherapy 2018; 73(6): 1700–7.

37. Sommer MO, Church GM, Dantas G. The human microbiome harbors a diverse reservoir of antibiotic resistance genes. Virulence 2010; 1(4): 299–303.

38. Baron SA, Diene SM, Rolain J-M. Human microbiomes and antibiotic resistance. Human Microbiome Journal 2018; 10: 43-52.

39. Corpet DE. Antibiotic resistance from food. The New England journal of medicine 1988; 318(18): 1206.

40. Mascaro V, Capano MS, Iona T, Nobile CGA, Ammendolia A, Pavia M. Prevalence of Staphylococcus aureus carriage and pattern of antibiotic resistance, including methicillin resistance, among contact sport athletes in Italy. Infection and drug resistance 2019; 12: 1161.

